# Intra-Ventricular Diastolic Hemodynamics under Pharmacological Stress in Patients with Ischemic Heart Disease: Analysis of 4D-Flow and Myocardial Flow Reserve using hybrid ^13^N-ammonia Positron Emission Tomography/Magnetic Resonance

**DOI:** 10.1101/2024.05.09.24307152

**Authors:** Keiichiro Endo, Kenji Fukushima, Masataka Katahira, Takatoyo Kiko, Ryo Yamakuni, Naoyuki Ukon, Takeshi Shimizu, Shiro Ishii, Takayoshi Yamaki, Kazuhiko Nakazato, Hiroshi Ito, Yasuchika Takeishi

**Author notes:** Corresponding author: Kenji Fukushima, M.D., Ph.D., Department of Radiology and Nuclear Medicine, Fukushima Medical University Hikarigaoka-1, Fukushima 960-1295, Japan.

## Abstract

**Objectives:** This study aimed to explore the linkage between intra-left ventricular (LV) diastolic hemodynamics and coronary endothelial function, utilizing four-dimensional (4D) flow magnetic resonance imaging (MR) and myocardial flow reserve (MFR) through simultaneous acquisition using hybrid PETMR system in patients with ischemic heart disease (IHD).

*Methods:* Sixty-eight patients (mean 66 ± 15 years, male 55) with IHD who underwent rest-pharmacological stress ^13^N-ammonia PET/MR were included. MFR and summed defect score (SSS and SRS for stress and rest) were obtained thorough rest-stress PET images. MR acquisition was performed simultaneously during PET scan to obtain rest-stress 4D flow datasets and followed by cine-MRI for the LV volume measurement. LV diastolic inflow(mL/s), peak velocity(cm/s), and averaged diastolic kinetic energy (KE)(μJ/mL) indexed with endo-diastolic volume were computed.

*Results:* Diastolic LV inflow parameters and KE significantly increased in stress scan compared to the rest (74.8 ± 17.5 cm/s vs. 64.5 ± 14.4 cm/s, p<0.0001; 10.1 ± 5.2 vs. 13.3 ± 7.8, p=0.0004 for peak velocity and KE, respectively). Stress KE showed a significant and weak correlation to MFR and SSS (r = 0.3, p=0.004; r=-0.4, p=0.002 for MFR and SSS, respectively). In patients with MFR above median value (1.76), stress KE significantly elevated from rest KE, while no significant change was observed for the patients with MFR below median (11.0 ± 4.6 vs. 16.2 ± 8.8, p=0.0002; 9.7 ± 5.4 vs. 10.3 ± 5.1 for rest vs. stress, respectively).

*Conclusion:* Non-invasive assessment of intra-LV diastolic hemodynamics derived from 4D flow MRI demonstrated significant alterations under stress, and was found to have a notable association with the extent of ischemia and coronary endothelial dysfunction.

## INTRODUCTION

Ischemic heart disease (IHD) is the major contributor to the global morbidity ^1^. Characterization of IHD by non-invasive imaging technique is a key to appropriate clinical decision making and early intervention. Myocardial perfusion imaging is the most routinely used technique in the evaluation of patients with IHD, allowing the detection of inducible myocardial ischemia and cardiac dysfunction. Myocardial blood flow (MBF), and myocardial flow reserve (MFR) derived from positron emission tomography (PET) provide useful information in the combination with the extent of perfusion defect and left ventricular (LV) function ^2,3^. Recently, LV diastolic dysfunction has been found to be associated with IHD. Several studies have shown the linkage between coronary atherosclerosis and the increased left ventricular stiffness ^4,5^. Impaired diastolic LV inflow dynamics has emerged as a potential biomarker, leading to structural remodeling and LV systolic dysfunction ^6^. Thus, in vivo quantification of diastolic LV inflow in three dimensions offers novel insights to assess the severity of ischemic burden. Echocardiography has been most used imaging tool to visualize diastolic LV inflow in clinical routine. However, the issues of insufficient penetration due to the limited acoustic window and the lack of reproducibility in the 2D visualization have been concerned ^7^. Cardiac magnetic resonance (MR) imaging has become more widely used tool to study myocardial tissue characterizations and cardiac function. In addition, four-dimensional (4D) flow MR imaging has been developed to simulate and quantify blood flow in vivo. Its utility has been initially validated for visualization and quantification of blood flow in great arteries, and the flow analysis using computational fluid dynamics provide novel hemodynamic markers such as the energy loss and the wall share stress ^8–10^. More recently, several studies have employed 4D flow MR to visualize and quantify intra LV blood flow to assess hemodynamic abnormality associated with various cardiac diseases including cardiomyopathy and structural heart disease, utilizing the quantification of the multi-directional blood stream computed as kinetic energy ^11,12^. However, the association with the extent of IHD, and coronary endothelial dysfunction has not been investigated. Furthermore, it remains unclear whether intra-LV flow alter to reflect the ischemia severity.

Hybrid PET/MR has emerged as an ideal modality in the evaluation of cardiac diseases, providing useful information on various pathological conditions through molecular and functional imaging in the combination of PET and MR as a one-stop examination ^13^. The aim of this study was to evaluate the usefulness of simultaneous analysis of PET perfusion and intra-LV 4D flow using hybrid ^13^N-ammonia PET/MR in patients with IHD.

## MATERIALS AND METHODS

### Patients

We retrospectively enrolled 76 consecutive patients known or suspect of IHD who underwent rest-pharmacological stress ^13^N-ammonia PET/MR from February 2022 to January 2024. Exclusion criteria were acute coronary syndrome, frequent arrhythmia including atrial fibrillation, implanted devices, symptomatic asthma, and pregnancy. We excluded patients who could not be included in the analysis because of poor image quality for 4D flow analysis. The study protocol was approved by the Ethics Committee at Fukushima Medical University and was conducted in accordance with the Declaration of Helsinki. All patients provided written informed consent before enrollment.

### 13N-ammonia PET/MR Imaging Protocol

PET and 4D flow MR were simultaneously conducted using a Biograph mMR (Siemens Healthineers, Erlangen, Germany) with integration of a single scanner. The PET component was built with avalanche photodiodes with lutetium oxyorthosilicate, with detectors that are not affected by magnetic fields from 3-Tesla scanner ^14^. The patients were instructed to refrain from any caffeine-containing products for 24 hours and requested to fast for more than 6 hours before the scan. Figure 1 shows rest-pharmacological stress ^13^N-ammonia PET/MRI protocol. To correct for PET attenuation of each patient in supine position, a two-point Dixon MRI sequence was performed at the beginning of the PET recording to obtain an attenuation map ^14,15^. After the attenuation correction sequence, the list mode PET scan and 4D-flow cine MR were simultaneously started. All ^13^N-ammonia PET images were acquired by 14-minute list-mode dynamic scan at rest and during vasodilator stress. We used continuous intravenous infusion of adenosine (160 µg/kg/minute), which was started 3 minutes before the stress scan until the end of the list-mode PET scan. A bolus of ^13^N-ammonium (500-700 MBq) was injected with saline flushes at intervals of 1 hour or longer between rest and the stress scan to obtain sufficient radioactive decay.

**Figure 1.**
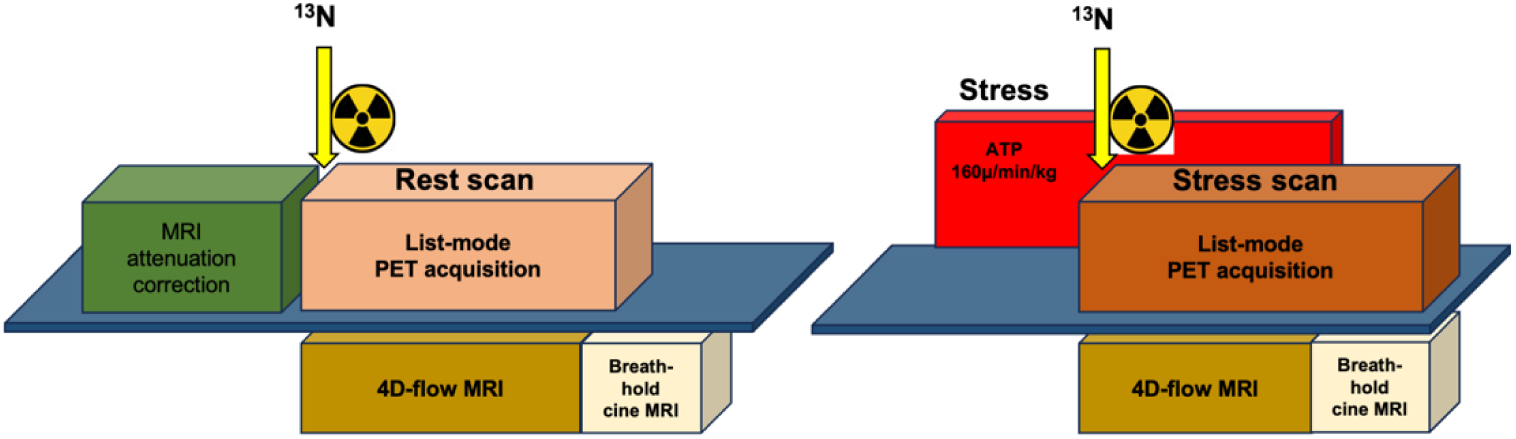
^13^N-Ammonia PET/MR acquisition protocol. After the localizer and attenuation map by MR was obtained, the list-mode PET acquisition and time-resolve, phase-contrast cine MR was simultaneously started. In stress scan, ATP (160 μg/min/kg for 10 min) was administrated 3 min prior and during PET/MR acquisition. 4D flow acquisition was followed by expiratory breath-hold, steady-state free cine MR was performed for rest and stress.

The 4D-flow MR acquisition protocol included electrocardiographic-gated, three-directional, time-resolved, phase-contrast cine sequences by free-breathing according to the latest statement ^16^. Scan parameters included: velocity encoding 1⁄4 200 cm/s for all directions, flip angle1⁄488, echo time1⁄43.7 ms, repetition time1⁄46.3 ms, parallel imaging (SENSE) speed-up factor1⁄42, and k-space segmentation factor 1⁄42. These settings gave a temporal resolution of 50.4 ms. The spatial resolution was 3 × 3 × 3 mm^3^. Velocity encoding was 150cm/s. No respiratory motion correction was used. The field-of-view was planned in rectangular form was adjusted for each subject to cover the left ventricle, atrium, and LV outflow in trans-axial plane. The repetition time was automatically adjusted according to the patient’s heart rate. Phase-contrast 4D cine was followed by end-expiratory breath-hold cine-MR. The standard long-axis views (two-chamber, three-chamber, and four-chamber views) and multiple short axis views were acquired, and the slice thickness was 5 mm. The number of phases obtained in each cardiac cycle was 25. The following parameters were applied: echo time of 1.5 ms, repetition time of 3.4 ms, matrix of 256×256 pixels, flip angle of 50° and typical field-of-view of 350 mm ^17^.

### PET image analysis

The imaging data obtained from list-mode PET acquisition were reconstructed using a three-dimensional attenuation-weighted ordered subset expectation maximization iterative reconstruction algorithm with 3 iterations and 21 subsets. The images were smoothed with a 2-mm full width at half maximum Gaussian filter ^13,14^. The image data matrix was 172×172, with a pixel size of 3.42 mm and a slice thickness of 2.03 mm. The presence and extent of perfusion impairment were quantified as summed stress score (SSS) and summed rest score (SRS) using a five-point scoring system and 17-segment model using commercially available, automatic calculation software based on ^82^Rb PET database (Corridor 4DM®, INVIA, MI). Summed difference score (SDS) was calculated as the difference between SSS and SRS. Dynamic data sets were analyzed by commercially available software (Syngo. via ver. 4. Siemens Healthineers, Erlangen, Germany) to obtain flow quantification MBF(mL/g/min) and MFR. A two-compartment model was used to quantify absolute MBF, which was calculated from PET images as previously described ^3^. The MFR was calculated as the ratio of hyperemic MBF to resting MBF.

### MRI image analysis

Postprocessing of the 4D-flow MR data was performed to be corrected for background errors and phase wraps to convert the data into a dicom format compatible with commercially available visualization software (GTF^®^ ver. 4.19, Gyro tools LLC, Sweden). Figure 2 shows the image analysis for intra-LV 4D flow. During the diastolic phase, the blood line passing through the mitral annuls was detected, indicating LV inflow (Figure 2A). The velocity magnitude map was created simultaneously, and highest magnitude (yellow to red colored) in early diastole was assigned for the region of interest. Using multi-oblique slices, the region of interest was placed on mitral annuls by semi-automatic contour tracing in perpendicular view to obtain the net flow volume (mL/s) and peak velocity (cm/s) of LV inflow. The inter-observer reproducibility for region of interest was assessed by two observers (MK and TK) for randomly assigned 20 patients.

**Figure 2.**
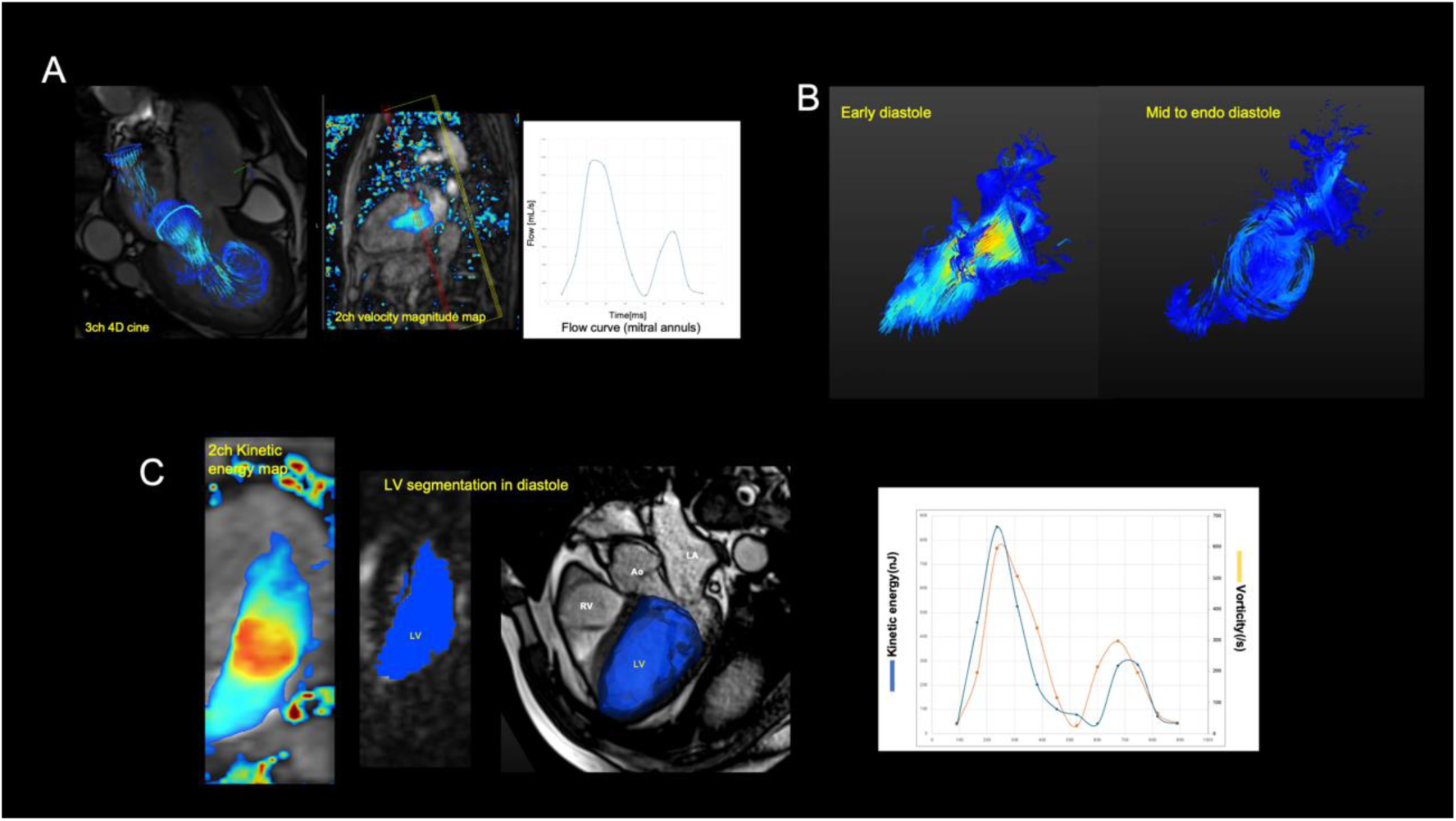
Representative images of intra-LV 4D flow analysis. LV inflow and magnitude velocity map during diastole are visualized, then the region of interest was placed on the area of mitral annuls by semi-automatic contour tracing with GTF® to obtain net LV inflow (mL/s) and peak velocity (cm/s) (A). The streamline of blood flow was visualized during cardiac cycle, and the phase of early systole to diastole was visually evaluated (B). In the end-diastole phase, LV was automatically segmented by using signal intensity threshold, then kinetic energy and vorticity map were created, and curves were obtained. Average and accumulated kinetic energy and vorticity indexed by end-diastolic volume in diastole phase was calculated (C, left). The peak E and A wave was obtained from the highest plot of diastole curve (C, right).

As shown in figure 2C, LV cavity in diastole phase was semi-automatically segmented using the region growing tool in GTF^®^, and the centerline for segmented cavity mask was automatically drawn by signal intensity through diastole to obtain LV mask at endo-diastole (Figure 2C left). Subsequently, kinetic energy (KE) and vorticity map was created to estimate energy volume passing through the mitral annulus and LV; the curve for averaged and accumulated KE, peak E to A wave, and vorticity through cardiac cycle were obtained and automatically calculated using GTF^®^ (Figure 2C right). Diastolic average KE, peak E-to A-wave, and vorticity were normalized by end-diastolic volume for rest and stress study as previously described ^9,11,18^. In visual analysis, abnormal flow was visualized as color-coded in stream-line images during early-to-mid diastole phase. LV volumetry measurement and curve analysis was conducted from cine-MRI to obtain LVEF, LVEDV, LVESV, peak filling rate (PFR)(/s) and time to filling rate for rest and stress using commercially available software (Segment®, Medviso, Lund, Sweden).

### Statistical Analysis

Data are presented as the mean ± standard deviation or number and percentage (%). Differences in continuous variables between two groups were compared using Student’s t-test. Categorical variables are expressed as counts and percentages and were compared by the χ^2^ test. Spearman’s correlation coefficient was used to assess the correlation between PET and 4D flow MR parameters. Comparison of parameters between rest and stress were performed using paired t test. Statistical analyses were performed using Prism^®^ version 9 (GraphPad software Inc. MA). P < 0.05 was considered statistically significant. The inter-class correlation was conducted to evaluated inter-observer reproducibility.

## RESULTS

### Patients’ demographics

Eight patients were excluded due to a failure of the 4D MR acquisition or due to insufficient data due to because of a rapid change in heart rate during the stress scan. Consequently, this study analyzed 136 images from 4D-flow scan. The patients’ demographics shown in Table 1. The mean age was 65.8 ± 15.1 years, and 51 (76.1%) patients were men. Fifty percent of the patients had a history of previous coronary interventions. More than half of the patients showed multi-vessel disease.

**Table 1.** Patients’ demographics.

| **<u>Characteristics</u>** |  |
| --- | --- |
| Age (years old) | 65.8 ± 15.1 |
| Male sex (n, %) | 51 (76.1) |
| Body mass index (kg/m<sup>2</sup>) | 24.2 ± 3.8 |
| **<u>Risk factors and past history</u>** |  |
| Hypertension (n, %) | 35 (52.2) |
| Diabetes mellitus (n, %) | 35 (52.2) |
| Dyslipidemia (n, %) | 52 (77.6) |
| Smoking history (n, %) | 39 (58.2) |
| Family history of CAD (n, %) | 15 (22.4) |
| Previous PCI (n, %) | 43 (64.2) |
| Previous CABG (n, %) | 8 (11.9) |
| Old myocardial infarction (n, %) | 21 (31.3) |
| Known CAD (n, %) |  |
| 1 vessel | 15 (22.4) |
| 2 vessels | 22 (32.8) |
| 3 vessels | 24 (35.8) |
| LMT | 3 (4.5) |

CAD, coronary artery disease; PCI, percutaneous coronary intervention; CABG, coronary artery bypass graft; LMT, left main trunk.

### LV 4D inflow dynamics and MFR

PET and MRI parameters are shown in Table 2. PET MBF was 0.7 ± 0.2 and 1.2 ± 0.4 ml/g/min for rest and stress, respectively. PET perfusion defect was 9.19 ± 8.31 (ranged 0 to 33) for SSS, and this was predominantly due to the defect in anterior wall. Table 3 shows PETMR parameters for rest and stress scan. LV volumetry, LVEF, and PFR showed a significant change in the stress scan from the rest. The intra-class correlation of region of interest was 0.75(95% coincidence interval 0.55 to 0.87). In the 4D-flow analysis for all patients, averaged KE was 0.1 ± 0.1 and 0.2 ± 0.09 (μJ/mL); averaged vorticity was 29.2 ± 8.1 and 32.5 ± 7.4 (/s) for rest and stress, respectively. Diastolic LV inflow parameters including stroke volume and peak velocity showed a significant increase in stress scan compared to the rest. Averaged KE and vorticity significantly increased in the stress scan compared to the rest. Figure 3 shows the correlations between 4D-flow and PETMR parameters. Average KE showed a significant moderate correlation with LVEF, and a weak correlation with PFR, MBF for both the rest and stress scan. Peak E wave showed a significant and moderate to strong correlation with PFR, while peak A wave showed mild correlation. Averaged vorticity showed a significant correlation to PETMR parameters including LVEF, PFR, MBF, and defect score in stress scan, while did not for the rest scan. Correlation coefficient, 95% coincidence interval, and p value are demonstrated in supplemental table. Figure 4 shows the representative plots for the correlations of KE, vorticity and PETMR parameters including MFR, SSS, and stress PFR. Averaged KE demonstrated a significant and weak correlation to MFR, SSS, and PFR (figure 4A, B, and C). Averaged vorticity showed a significant and weak correlation to MFR (figure 4D). Table 4 shows 4D flow and PET perfusion defect. Volumetry showed a significant change in abnormal perfusion for rest and stress (SSS≥4, SRS≥4), while remained unchanged for normal and abnormal stress perfusion. Average KE showed a significantly increased among the patients with normal perfusion and abnormal stress perfusion, while remained unchanged in patients with abnormal perfusion for rest and stress. In addition, as shown in table 5 and figure 5, when the patients were divided above or below median (MFR≥1.76, n=34; MFR<1.76, n=34), averaged KE showed a significant increase in stress scan compared to the rest (11.0 ± 4.6 vs. 16.2 ± 8.8, p=0.0002; 9.7 ± 5.4 vs. 10.3 ± 5.1 for rest vs. stress, respectively), while did not for the patients with MFR below median. Figure 5 shows two representative cases for normal and abnormal LV inflow during stress. A demonstrates normal perfusion and cardiac function with preserved global MFR. The averaged KE was 13.2 μJ /mL for rest scan and elevated to 37.1 μJ /mL during stress scan. B illustrates a patient with a significant anterior infarction and ischemia with reduced cardiac function and MFR. A retained, vortex flow was observed along mid anterior wall during diastole (figure 5B, red arrow). Averaged KE was 16.7μJ /mL for rest, and reduced to 14.3μJ /mL under stress.

**Figure 3.**
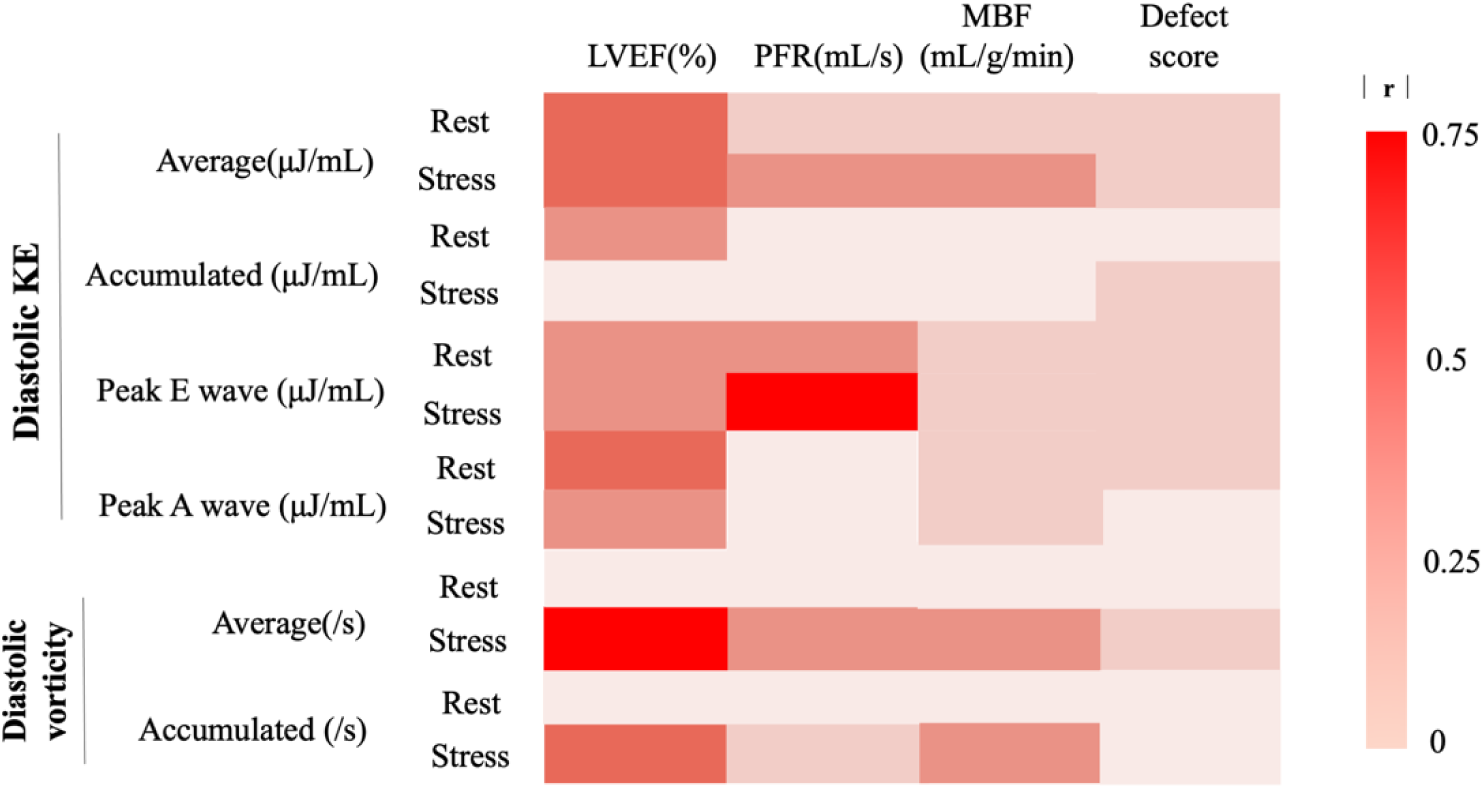
Correlation matrix between 4D flow and PETMR parameters. The color scale represents the correlation between two variables as the absolute r value. Red highlights strong correlation while pink highlights weak correlation. Average KE showed a significant moderate correlation with LVEF for rest and stress scan, and a weak to mild correlation with PFR, MBF for stress, while not significant for rest scan. Peak E-wave showed a significant and moderate to strong correlation with PFR, while peak A-wave showed mild correlation. Averaged vorticity showed a significant correlation to LVEF, PFR, MBF, and defect score in stress scan, while did not for the rest scan.

**Figure 4.**
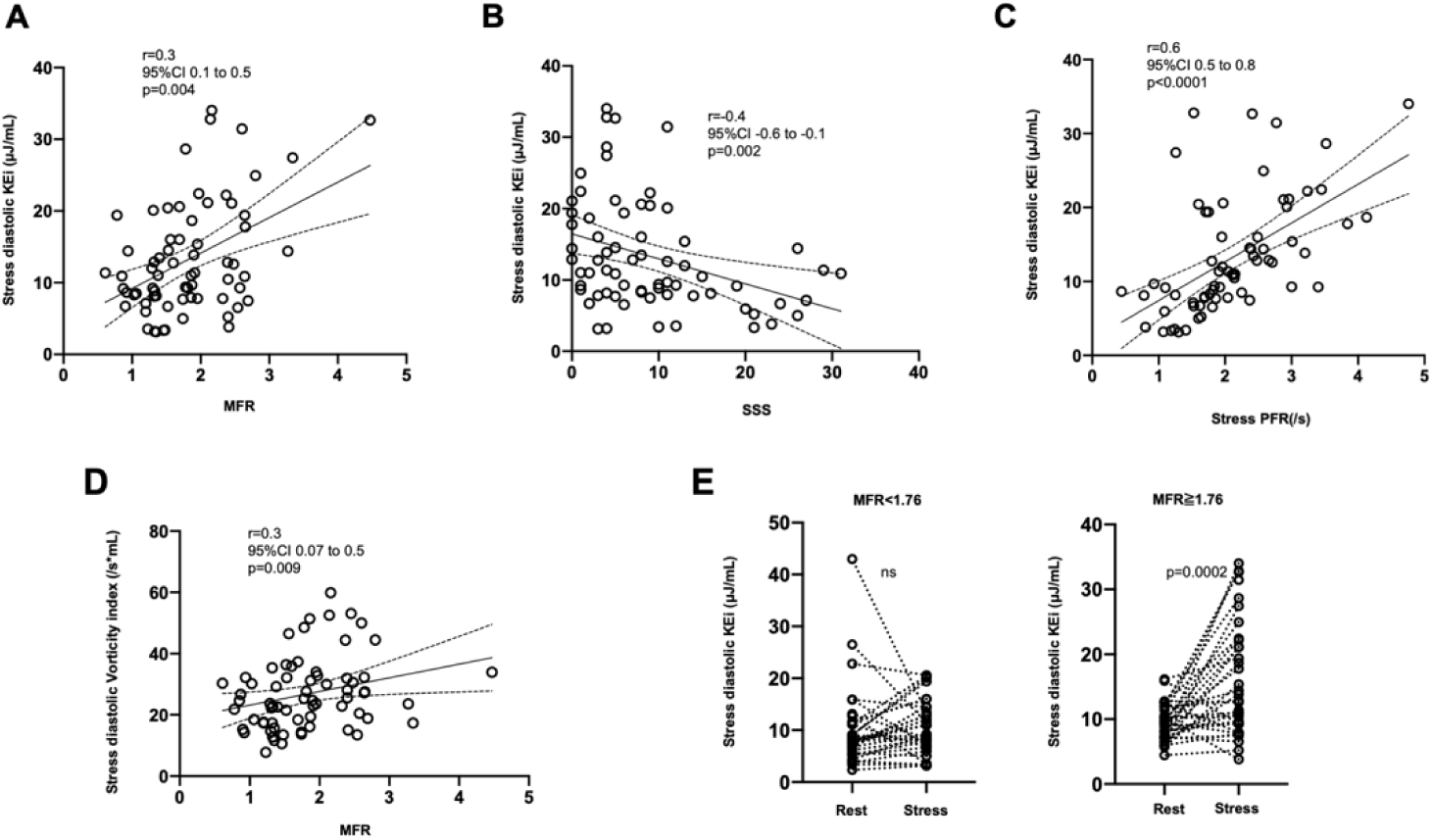
The scatter plots illustrating the relationship between PETMR and 4D flow MR parameters. Stress KE showed a significant correlation to global MFR, SSS, and PFR(A, B, and C). Stress vorticity showed a significant and weak correlation to MFR. E showed comparison between rest stress for the patients with MFR above or below median value. Averaged KE showed a significant increase in the stress compared to the rest in patients with MFR above median (right), while no significant change was observed for the patients with MFR below median (left).

**Figure 5.**
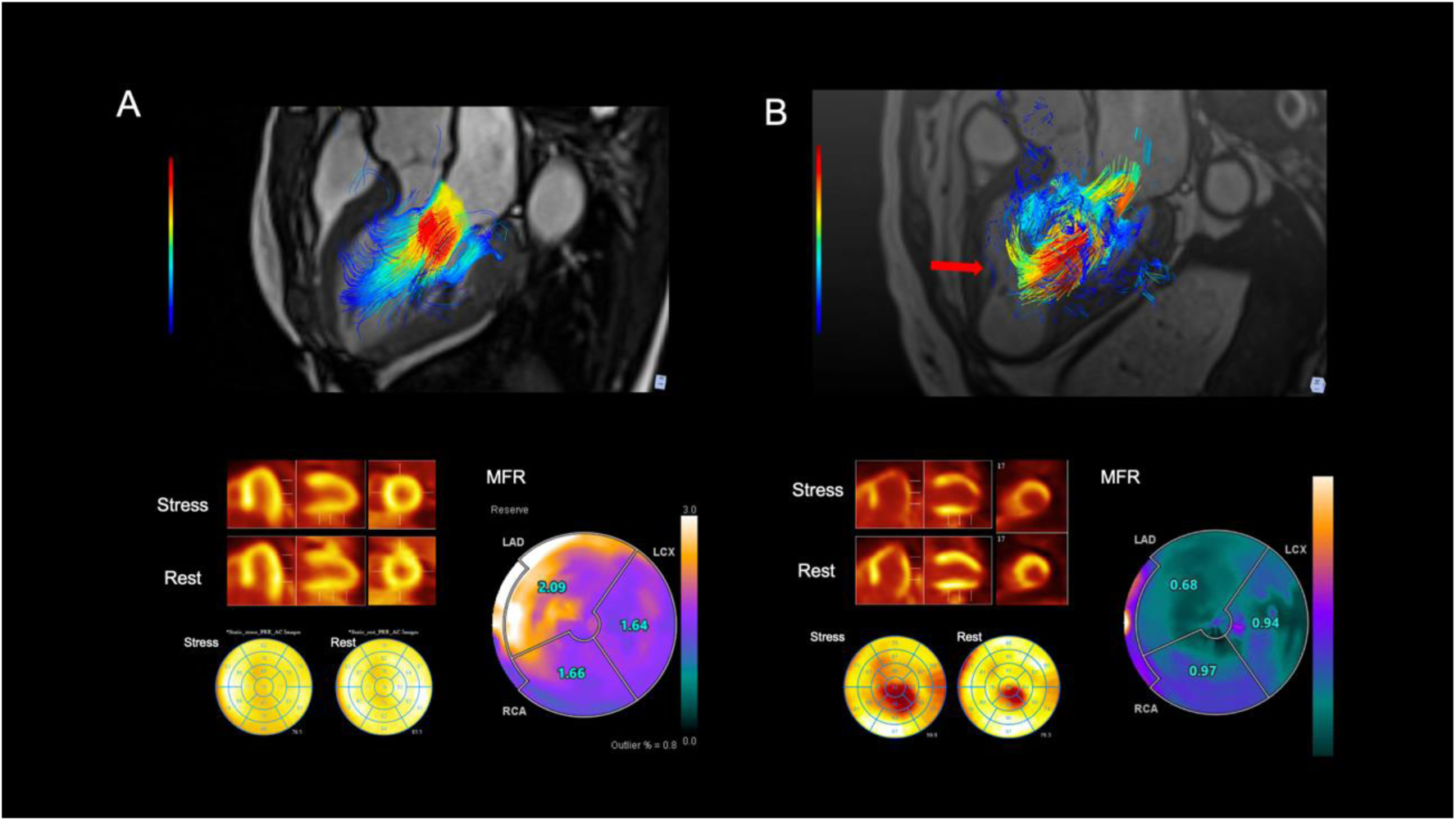
Representative cases for normal and abnormal LV inflow are shown. A was for normal perfusion and cardiac function with preserved MFR in LAD territory (SSS=0, stress LVEF 50%, and MFR 1.9), demonstrating normal LV inflow velocity and averaged KE (65cm/s, and 13.2μ J/mL for rest; 85cm/s, and 37.1μJ/mL for stress). B was for a patient with a significant anterior infarction and ischemia with impaired cardiac function and MFR (SSS=24, stress LVEF 35%, and MFR 0.8). The pass line visualization demonstrated a retained and vortex flow in mid anterior during mid-diastole phase (red arrow). LV inflow and averaged KE were 59cm/s, and 16.7μJ/mL for rest; 60cm/s, and 14.3μJ/mL for stress.

**Table 2.** PET parameters (all patients)

|  |  |
| --- | --- |
| **<u>MBF</u>** |  |
| Global stress (mL/g/min) | 1.2 ± 0.5 |
| Global rest (mL/g/min) | 0.7 ± 0.2 |
|  |  |
| ***Global MFR*** | 1.8 ± 0.7 |
| LAD-MFR | 1.8 ± 0.7 |
| LCX-MFR | 1.9 ± 0.7 |
| RCA-MFR | 2.0 ± 0.8 |
|  |  |
| **<u>Perfusion defect</u>** |  |
| Summed stress score (SSS) | 9.2 ± 8.3 |
| LAD-SSS | 6.3 ± 6.7 |
| LCX-SSS | 1.7 ± 6.7 |
| RCA-SSS | 1.2 ± 2.2 |
|  |  |
| Summed rest score (SRS) | 4.5 ± 5.9 |
| Summed differential score (SDS) | 4.8 ± 5.7 |

MBF myocardial blood flow, MFR myocardial flow reserve, LAD left anterior descending artery, LCX left circumferential artery, RCA right coronary artery, SSS summed stress score, SRS summed rest score, SDS summed differential score

**Table 3.** MRI parameters (all patients)

|  | Rest | Stress | *p* |
| --- | --- | --- | --- |
| ***Cine-MRI*** |  |  |  |
| EDV(mL) | 127.3 ± 54.1 | 143.7 ± 61.2 | 0.003 |
| ESV(mL) | 68.9 ± 50.7 | 77.1 ± 57.7 | 0.001 |
| LVEF(%) | 51.3 ± 15.9 | 51.8 ± 16.3 | 0.6 |
| SV(mL) | 58.4 ± 13.4 | 66.7 ± 16.4 | <0.0001 |
| PFR(mL/s) | 1.9 ± 0.7 | 2.1 ± 0.8 | 0.004 |
| ***4D-Flow MRI*** |  |  |  |
| <u>LV outflow</u> |  |  |  |
| Net stroke volume(mL/s) | 38.3 ± 28.1 | 44.8 ± 32.4 | 0.03 |
| Peak Velocity(cm/s) | 80.4 ± 20.6 | 83.6 ± 24.7 | 0.2 |
| <u>LV inflow</u> |  |  |  |
| Net stroke volume(mL/s) | 54.0 ± 22.5 | 69.4 ± 31.4 | <0.0001 |
| Average velocity (mL/s) | 74.9 ± 35.3 | 55.7 ± 22.2 | <0.0001 |
| Peak Velocity(cm/s) | 64.5 ± 14.4 | 74.8 ± 17.5 | <0.0001 |
| <u>Diastolic KE</u> |  |  |  |
| Average(μJ) | 115.6 ± 60.6 | 162.1 ± 77.0 | 0.02 |
| EDVindexed(μJ/mL) | 10.1 ± 5.2 | 13.3 ± 7.8 | 0.0004 |
| Accumulated(μJ) | 1665.6 ± 723.6 | 1918.3 ± 994.8 | 0.04 |
| EDVindexed(μJ/mL) | 145.9 ± 77.9 | 148.6 ± 74.4 | 1.0 |
| Peak(μJ) | 9970.8 ± 5395.3 | 9084.7 ± 5148.9 | 0.2 |
| EDVindexed(μJ/mL) | 90.5 ± 53.7 | 77.1 ± 45.8 | 0.3 |
| Peak E wave<br>(EDVindexed(μJ/mL)) | 25.2 ± 24.3 | 28.1 ± 20.4 | 0.1 |
| Peak A wave<br>(EDVindexed(μJ/mL)) | 20.9 ± 18.3 | 27.1 ± 19.3 | 0.07 |
| <u>Diastolic Vorticity</u> |  |  |  |
| Average(/s) | 29.2 ± 8.1 | 32.5 ± 7.7 | 0.0005 |
| Accumulated(/s) | 31736.6 ± 19224.6 | 29519.9 ± 16345.7 | 0.1 |

EDV, left ventricular end-diastolic volume; ESV, left ventricular end-systolic volume; LVEF, left ventricular ejection fraction; SV stroke volume; PFR, peak filling rate, KE kinetic energy

**Table 4.** PET perfusion and MR parameters.

|  | Normal perfusion<br>(SSS≤3, SRS≤3)(n=17) |  |  | Abnormal perfusion |  |  |  |  |  |
| --- | --- | --- | --- | --- | --- | --- | --- | --- | --- |
|  |  |  |  | (SSS≥4, SRS≤3)(n=24) |  |  | (SSS≥4, SRS≥4)(n=27) |  |  |
|  | Rest | Stress | p | Rest | Stress | p | Rest | Stress | p |
| **Cine-MR parameter** |  |  |  |  |  |  |  |  |  |
| LVEDV(mL) | 113.8 ± 31.1 | 113.6 ± 26.5 | 1.0 | 131.4 ± 27.9 | 121.8 ± 25.1 | 0.1 | 150.7 ± 63.7 | 172.9 ± 74.9 | <0.0001 |
| LVESV(mL) | 53.3 ± 28.2 | 48.6 ± 25.7 | 0.2 | 64.6 ± 30.1 | 55.5 ± 17.5 | 0.2 | 91.8 ± 60.1 | 106.1 ± 70.4 | 0.008 |
| LVEF(%) | 55.1 ± 12.7 | 58.7 ± 12.1 | 0.09 | 52.8 ± 9.3 | 54.7 ± 11.1 | 0.4 | 44.5 ± 16.4 | 43.9 ± 16.0 | 0.7 |
| SV(mL) | 60.5 ± 16.0 | 65.1 ± 13.2 | 0.07 | 66.8 ± 16.1 | 66.4 ± 17.0 | 0.9 | 58.9 ± 16.1 | 66.8 ± 18.7 | 0.0002 |
| PFR(/s) | 2.1 ± 0.7 | 2.3 ± 0.9 | 0.3 | 2.0 ± 0.8 | 2.2 ± 0.8 | 0.4 | 1.7 ± 0.7 | 1.9 ± 0.8 | 0.08 |
| **4D-flow parameter** |  |  |  |  |  |  |  |  |  |
| LV-inflow net volume(mL/s) | 52.2 ± 27.2 | 70.7 ± 34.2 | 0.01 | 57.9 ± 21.4 | 74.3 ± 29.4 | 0.002 | 51.8 ± 20.9 | 64.6 ± 31.9 | 0.002 |
| LV-inflow peak velocity(cm/s) | 58.4 ± 14.8 | 74.8 ± 14.0 | 0.01 | 65.8 ± 15.8 | 76.0 ± 16.2 | 0.03 | 66.9 ± 12.5 | 73.7 ± 20.6 | 0.02 |
| LV-outflow net volume(mL/s) | 40.6 ± 29.5 | 50.6 ± 31.8 | 0.09 | 32.3 ± 21.2 | 38.9 ± 27.4 | 0.2 | 42.0 ± 32.0 | 46.1 ± 37.2 | 0.5 |
| LV-outflow peak velocity(cm/s) | 81.0 ± 21.2 | 84.0 ± 25.9 | 0.4 | 78.8 ± 14.4 | 83.3 ± 18.7 | 0.2 | 81.4 ± 24.7 | 83.5 ± 29.3 | 0.7 |
| Average KEi(μJ/mL) | 10.7 ± 3.3 | 14.0 ± 6.1 | 0.03 | 9.6 ± 4.1 | 15.1 ± 8.4 | 0.001 | 10.3 ± 6.0 | 11.2 ± 7.9 | 0.5 |
| Peak E wave | 35.2 ± 42.8 | 36.4 ± 21.5 | 0.8 | 45.5 ± 20.2 | 29.4 ± 22.0 | 0.001 | 21.5 ± 14.1 | 21.7 ± 16.6 | 0.9 |
| Peak A wave | 22.0 ± 22.4 | 23.9 ± 21.6 | 0.1 | 15.8 ± 11.1 | 23.4 ± 17.9 | 0.04 | 13.1 ± 8.0 | 16.1 ± 15.5 | 0.4 |
| Diastolic Vorticity(/s) | 28.2 ± 10.8 | 29.1 ± 12.6 | 0.7 | 28.5 ± 12.7 | 27.9 ± 11.5 | 0.4 | 24.0 ± 14.3 | 24.1 ± 11.7 | 0.9 |

EDV, left ventricular end-diastolic volume; ESV, left ventricular end-systolic volume; LVEF, left ventricular ejection fraction; SV stroke volume; PFR, peak filling rate, LV left ventricular, KE kinetic energy, Kei kinetic energy EDV indexed

**Table 5.** Flow reserve and MR parameters.

| | MFR≥1.76 (n=34) | | | MFR<1.76 (n=34) | | |
| --- | --- | --- | --- | --- | --- | --- |
|  | Rest | Stress | p value | Rest | Stress | p value |
| **<u>Cine-MR parameter</u>** |  |  |  |  |  |  |
| LVEDV(mL) | 126.7 ± 34.6 | 118.5 ± 30.2 | 0.1 | 144.4 ± 44.7 | 118.5 ± 30.2 | 0.1 |
| LVESV(mL) | 66.3 ± 39.9 | 51.6 ± 26.6 | 0.005 | 83.6 ± 44.3 | 51.6 ± 26.6 | 0.05 |
| LVEF(%) | 50.8 ± 13.7 | 57.7 ± 11.6 | 0.0006 | 44.9 ± 15.0 | 57.7 ± 11.6 | 0.07 |
| SV(mL) | 60.0 ± 16.1 | 66.9 ± 15.2 | 0.03 | 60.8 ± 39.8 | 66.9 ± 15.2 | 1.0 |
| PFR(/s) | 2.1 ± 0.8 | 2.5 ± 0.9 | 0.008 | 1.7 ± 0.6 | 2.5 ± 0.9 | 1.0 |
| **<u>4D-flow parameter</u>** |  |  |  |  |  |  |
| LV-inflow net volume(mL/s) | 54.2 ± 19.8 | 68.5 ± 30.1 | 0.0009 | 53.7 ± 25.1 | 70.4 ± 33.1 | 0.0001 |
| LV-inflow peak velocity(cm/s) | 65.0 ± 15.1 | 75.6 ± 17.7 | 0.01 | 64.0 ± 14.0 | 73.9 ± 17.4 | 0.001 |
| Average KE(μJ/mL) | 11.0 ± 4.6 | 16.2 ± 8.8 | 0.0007 | 9.7 ± 5.4 | 10.3 ± 5.1 | 0.3 |
| Peak E wave (μJ/mL) | 29.1 ± 31.6 | 32.7 ± 20.8 | 0.4 | 23.0 ± 13.7 | 22.4 ± 14.3 | 0.9 |
| Peak A wave (μJ/mL) | 19.3 ± 17.7 | 23.9 ± 20.7 | 0.05 | 13.0 ± 8.0 | 16.3 ± 11.5 | 0.1 |
| Average Vorticity(/s/mL) | 28.7 ± 10.5 | 31.2 ± 12.3 | 0.1 | 0.2 ± 0.1 | 0.2 ± 0.1 | 0.2 |

EDV, left ventricular end-diastolic volume; ESV, left ventricular end-systolic volume; LVEF, left ventricular ejection fraction; SV stroke volume; PFR, peak filling rate, LV left ventricular, KE kinetic energy, KEi kinetic energy EDV indexed

## DISCUSSION

In the present study, we conducted a simultaneous evaluation of intra-LV diastolic flow and coronary endothelial function using ^13^N-ammonia PET/MR to explore the association with the extent of IHD. MR-derived mitral flow significantly correlated with MFR. LV-inflow parameters and diastolic average KE demonstrated a significant association with established diastolic filling parameters, MFR, and the extent of perfusion defect. Furthermore, KE was altered during pharmacological stress and significantly increased in patients with preserved MFR. The intra-LV 4D flow analysis including healthy controls has been extensively validated ^8,19^. Numerous studies have confirmed that intra-LV blood flow is closely related to the LV morphology, wall kinetics, and diastolic compliance in patients with heart failure and previous myocardial infarction, suggesting that 4D-flow CMR is the reference for non-invasive assessment of intra-LV blood flow hemodynamics ^8,9,11,19,20^. These initial studies reported that the average KE in early diastole was significantly increased in patients with disease such as dilated cardiomyopathy or myocardial infarction compared to healthy controls ^11,21^. However, the findings of the present study appear to challenge these earlier reports. Nonetheless, increased KE has been initially observed as turbulent flow in outflow obstruction ^20^. Stoll et al. reported that diastolic KE was significantly decreased in patients with heart failure compared to healthy controls ^22^. Additionally, Das et al. reported that KE was associated with infarct size, showing a trend to decrease in the group with severe infarction ^23^. Furthermore, in a study enrolled healthy controls and athletes, increased diastolic KE was observed in athletes, with a decrease noted with age ^19,24^. Thus, the ongoing debate regarding the alteration of KE remains unresolved. Technically, phase-contrast-derived 4D-flow MR consists of encoded velocity datasets for blood flow in multi-directions which pass through mitral annuls considered as the narrowest waypoint in diastole. KE typically represents the energy associated with the blood flow and mass at each point during the cardiac cycle, simulating turbulent flow. The results of our study indicated that LV inflow markedly increase reflecting transient hyperdynamic wall motion due to vasodilation, namely intra LV flow may alter and produce rapid vortex flow, resulting increased KE values. The same explanation can be applied to the increased vorticity observed under stress. Since the PET parameters have been established as robust markers for the extent of IHD, the result of our study indicated that the impaired coronary endothelial function decreases myocardial relaxation during diastolic phase, causing increased diastolic pressure. Possible reason of insufficient increase of KE under stress is that the LV dilation predominantly consists long-axial component, namely LV inflow is primarily driven by longitudinal force from the point of mitral annuls to distal apex. Myocardial ischemia may increase diastolic myocardial stiffness and weaken longitudinal force of inflow passing through the mitral annuls. Previous studies by Ben-Arzi et al. revealed that increased wall stress caused by intra-LV blood flow hemodynamics abnormality was a powerful stimulant for the sympathetic adrenergic system ^18^. Garg et al. demonstrated that diastolic KE decreases after myocardial infarction even in patients with preserved LV ejection fraction ^25^. This suggests that diastolic dysfunction may serve as an early sign in the temporal sequence of myocardial ischemia in IHD, preceding overt systolic dysfunction. The results of our study revealed the potential strength in simultaneous non-invasive assessment of LV inflow parameters during stress in the combination with established markers.

This study has some limitations. MR acquisition is substantially influenced by heart rate variability. Therefore, in this study, a continuous infusion of adenosine during acquisition was employed, and field of view was set to cover LV by excluding the great vessels to shorten scan duration. Despite these efforts, the insufficient 4D scans were obtained among excluded patients. The results of this study are not fully applicable to patients with significant valvular disease, and frequent arrhythmia. In our study, the particle analysis of flow components was not available. We analyzed a small number of patients from a single center, and reproducibility among MR scanner and software needs to be validated. Further studies with a larger sample size are warranted. Nevertheless, this is the first study to report the simultaneous assessment of MFR and 4D flow MRI through a single imaging session with rest-stress ^13^N-ammonia PET/MRI, which has a potential to serve as a novel indicator for assessing the severity of ischemia in patients with IHD.

## CONCLUSION

Intra-LV 4D flow dynamics was altered under pharmacological stress and associated with coronary endothelial function in patients with IHD. The simultaneous analysis of PET perfusion and 4D-flow using hybrid PET/MR system has a potential role to serve as a novel imaging technique for the hemodynamic assessment and the extent of ischemia.

## Data Availability

not available

## DISCLOSURE

KF was supported by JSPS-KAKENHI (grant No. 22K07701) Other authors declare no conflict of interest.

---

**Supplemental Table.**
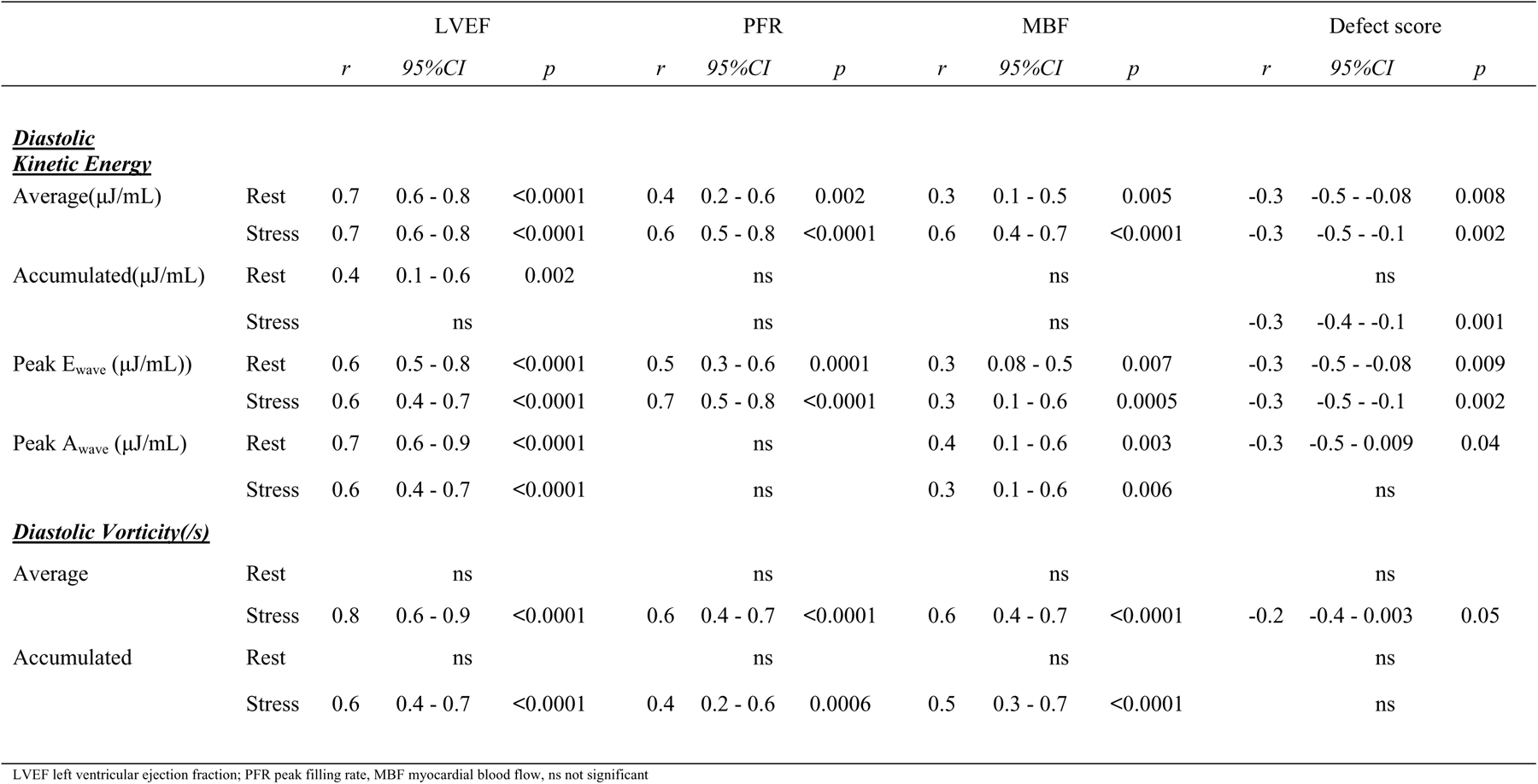
Correlations between 4D-flow and PETMR parameters.

|  |  | LVEF |  |  | PFR |  |  | MBF |  |  | Defect score |  |  |
| --- | --- | --- | --- | --- | --- | --- | --- | --- | --- | --- | --- | --- | --- |
|  |  | *r* | *95%CI* | *p* | *r* | *95%CI* | *p* | *r* | *95%CI* | *p* | *r* | *95%CI* | *p* |
| <u>**Diastolic Kinetic Energy**</u> |  |  |  |  |  |  |  |  |  |  |  |  |  |
| Average(μJ/mL) | Rest | 0.7 | 0.6 - 0.8 | <0.0001 | 0.4 | 0.2 - 0.6 | 0.002 | 0.3 | 0.1 - 0.5 | 0.005 | -0.3 | -0.5 - -0.08 | 0.008 |
|  | Stress | 0.7 | 0.6 - 0.8 | <0.0001 | 0.6 | 0.5 - 0.8 | <0.0001 | 0.6 | 0.4 - 0.7 | <0.0001 | -0.3 | -0.5 - -0.1 | 0.002 |
| Accumulated(μJ/mL) | Rest | 0.4 | 0.1 - 0.6 | 0.002 | ns |  |  | ns |  |  | ns |  |  |
|  | Stress | ns |  |  | ns |  |  | ns |  |  | -0.3 | -0.4 - -0.1 | 0.001 |
| Peak E<sub>wave</sub> (μJ/mL)) | Rest | 0.6 | 0.5 - 0.8 | <0.0001 | 0.5 | 0.3 - 0.6 | 0.0001 | 0.3 | 0.08 - 0.5 | 0.007 | -0.3 | -0.5 - -0.08 | 0.009 |
|  | Stress | 0.6 | 0.4 - 0.7 | <0.0001 | 0.7 | 0.5 - 0.8 | <0.0001 | 0.3 | 0.1 - 0.6 | 0.0005 | -0.3 | -0.5 - -0.1 | 0.002 |
| Peak A<sub>wave</sub> (μJ/mL) | Rest | 0.7 | 0.6 - 0.9 | <0.0001 | ns |  |  | 0.4 | 0.1 - 0.6 | 0.003 | -0.3 | -0.5 - 0.009 | 0.04 |
|  | Stress | 0.6 | 0.4 - 0.7 | <0.0001 | ns |  |  | 0.3 | 0.1 - 0.6 | 0.006 | ns |  |  |
| <u>**Diastolic Vorticity(/s)**</u> |  |  |  |  |  |  |  |  |  |  |  |  |  |
| Average | Rest | ns |  |  | ns |  |  | ns |  |  | ns |  |  |
|  | Stress | 0.8 | 0.6 - 0.9 | <0.0001 | 0.6 | 0.4 - 0.7 | <0.0001 | 0.6 | 0.4 - 0.7 | <0.0001 | -0.2 | -0.4 - 0.003 | 0.05 |
| Accumulated | Rest | ns |  |  | ns |  |  | ns |  |  | ns |  |  |
|  | Stress | 0.6 | 0.4 - 0.7 | <0.0001 | 0.4 | 0.2 - 0.6 | 0.0006 | 0.5 | 0.3 - 0.7 | <0.0001 | ns |  |  |

LVEF left ventricular ejection fraction; PFR peak filling rate, MBF myocardial blood flow, ns not significant

## Notes

### Competing Interest Statement

The authors have declared no competing interest.

### Author Declarations

The Ethics Committee at Fukushima Medical University

## References

1. Nowbar AN, Gitto M, Howard JP, Francis DP, Al-Lamee R. Mortality From Ischemic Heart Disease. Circulation: Cardiovascular Quality and Outcomes. 2019;12:e005375. doi: 10.1161/CIRCOUTCOMES.118.005375

2. Schindler Thomas H, Fearon William F, Pelletier-Galarneau M, Ambrosio G, Sechtem U, Ruddy Terrence D, Patel Krishna K, Bhatt Deepak L, Bateman Timothy M, Gewirtz H, et al. Myocardial Perfusion PET for the Detection and Reporting of Coronary Microvascular Dysfunction. JACC: Cardiovascular Imaging. 2023;16:536–548. doi: 10.1016/j.jcmg.2022.12.015

3. Ziadi MC, Dekemp RA, Williams KA, Guo A, Chow BJ, Renaud JM, Ruddy TD, Sarveswaran N, Tee RE, Beanlands RS. Impaired myocardial flow reserve on rubidium-82 positron emission tomography imaging predicts adverse outcomes in patients assessed for myocardial ischemia. J Am Coll Cardiol. 2011;58:740–748. doi: 10.1016/j.jacc.2011.01.065

4. Aggarwal P, Sinha SK, Marwah R, Nath RK, Pandit BN, Singh AP. Effect of Percutaneous Coronary Intervention on Diastolic Function in Coronary Artery Disease. J Cardiovasc Echogr. 2021;31:73–76. doi: 10.4103/jcecho.jcecho_128_20

5. Lv L, Ma X, Xu Y, Zhang Q, Kan S, Chen X, Liu H, Wang H, Wang C, Ma J. The constricting effect of reduced coronary artery compliance on the left ventricle is an important cause of reduced diastolic function in patients with coronary heart disease. BMC Cardiovascular Disorders. 2022;22:375. doi: 10.1186/s12872-022-02809-0

6. Zile MR, Brutsaert DL. New Concepts in Diastolic Dysfunction and Diastolic Heart Failure: Part I. Circulation. 2002;105:1387–1393. doi: 10.1161/hc1102.105289

7. Nagueh SF. Left Ventricular Diastolic Function: Understanding Pathophysiology, Diagnosis, and Prognosis With Echocardiography. JACC: Cardiovascular Imaging. 2020;13:228–244. doi: 10.1016/j.jcmg.2018.10.038

8. Crandon S, Elbaz MSM, Westenberg JJM, van der Geest RJ, Plein S, Garg P. Clinical applications of intra-cardiac four-dimensional flow cardiovascular magnetic resonance: A systematic review. Int J Cardiol. 2017;249:486–493. doi: 10.1016/j.ijcard.2017.07.023

9. Demirkiran A, Hassell MECJ, Garg P, Elbaz MSM, Delewi R, Greenwood JP, Piek JJ, Plein S, van der Geest RJ, Nijveldt R. Left ventricular four-dimensional blood flow distribution, energetics, and vorticity in chronic myocardial infarction patients with/without left ventricular thrombus. European Journal of Radiology. 2022;150:110233. doi: 10.1016/j.ejrad.2022.110233

10. Grafton-Clarke C, Crandon S, Westenberg JJM, Swoboda PP, Greenwood JP, van der Geest RJ, Swift AJ, Vassiliou VS, Plein S, Garg P. Reproducibility of left ventricular blood flow kinetic energy measured by four-dimensional flow CMR. BMC Research Notes. 2021;14:289. doi: 10.1186/s13104-021-05697-3

11. Eriksson J, Bolger AF, Ebbers T, Carlhäll CJ. Four-dimensional blood flow-specific markers of LV dysfunction in dilated cardiomyopathy. Eur Heart J Cardiovasc Imaging. 2013;14:417–424. doi: 10.1093/ehjci/jes159

12. Eriksson J, Bolger AF, Ebbers T, Carlhäll CJ. Assessment of left ventricular hemodynamic forces in healthy subjects and patients with dilated cardiomyopathy using 4D flow MRI. Physiol Rep. 2016;4. doi: 10.14814/phy2.12685

13. Rischpler C, Nekolla SG, Kunze KP, Schwaiger M. PET/MRI of the heart. Semin Nucl Med. 2015;45:234–247. doi: 10.1053/j.semnuclmed.2014.12.004

14. Masuda A, Nemoto A, Takeishi Y. Technical aspects of cardiac PET/MRI. J Nucl Cardiol. 2018;25:1023–1028. doi: 10.1007/s12350-018-1237-4

15. Kiko T, Yokokawa T, Misaka T, Masuda A, Yoshihisa A, Yamaki T, Kunii H, Nakazato K, Takeishi Y. Myocardial viability with chronic total occlusion assessed by hybrid positron emission tomography/magnetic resonance imaging. J Nucl Cardiol. 2021;28:2335–2342. doi: 10.1007/s12350-020-02041-3

16. Bissell MM, Raimondi F, Ait Ali L, Allen BD, Barker AJ, Bolger A, Burris N, Carhäll C-J, Collins JD, Ebbers T, et al. 4D Flow cardiovascular magnetic resonance consensus statement: 2023 update. Journal of Cardiovascular Magnetic Resonance. 2023;25:40. doi: 10.1186/s12968-023-00942-z

17. Schulz-Menger J, Bluemke DA, Bremerich J, Flamm SD, Fogel MA, Friedrich MG, Kim RJ, von Knobelsdorff-Brenkenhoff F, Kramer CM, Pennell DJ, et al. Standardized image interpretation and post-processing in cardiovascular magnetic resonance - 2020 update : Society for Cardiovascular Magnetic Resonance (SCMR): Board of Trustees Task Force on Standardized Post-Processing. J Cardiovasc Magn Reson. 2020;22:19. doi: 10.1186/s12968-020-00610-6

18. Ben-Arzi H, Das A, Kelly C, van der Geest RJ, Plein S, Dall’Armellina E. Longitudinal Changes in Left Ventricular Blood Flow Kinetic Energy After Myocardial Infarction: Predictive Relevance for Cardiac Remodeling. J Magn Reson Imaging. 2022;56:768–778. doi: 10.1002/jmri.28015

19. Crandon S, Westenberg JJM, Swoboda PP, Fent GJ, Foley JRJ, Chew PG, Brown LAE, Saunderson C, Al-Mohammad A, Greenwood JP, et al. Impact of Age and Diastolic Function on Novel, 4D flow CMR Biomarkers of Left Ventricular Blood Flow Kinetic Energy. Scientific Reports. 2018;8:14436. doi: 10.1038/s41598-018-32707-5

20. Demirkiran A, van Ooij P, Westenberg JJM, Hofman MBM, van Assen HC, Schoonmade LJ, Asim U, Blanken CPS, Nederveen AJ, van Rossum AC, et al. Clinical intra-cardiac 4D flow CMR: acquisition, analysis, and clinical applications. Eur Heart J Cardiovasc Imaging. 2022;23:154–165. doi: 10.1093/ehjci/jeab112

21. Corrado PA, Macdonald JA, François CJ, Aggarwal NR, Weinsaft JW, Wieben O. Reduced regional flow in the left ventricle after anterior acute myocardial infarction: a case control study using 4D flow MRI. BMC Medical Imaging. 2019;19:101. doi: 10.1186/s12880-019-0404-7

22. Stoll VM, Hess AT, Rodgers CT, Bissell MM, Dyverfeldt P, Ebbers T, Myerson SG, Carlhäll C-J, Neubauer S. Left Ventricular Flow Analysis. Circulation: Cardiovascular Imaging. 2019;12:e008130. doi: 10.1161/CIRCIMAGING.118.008130

23. Das A, Kelly C, Ben-Arzi H, van der Geest RJ, Plein S, Dall’Armellina E. Acute intra-cavity 4D flow cardiovascular magnetic resonance predicts long-term adverse remodelling following ST-elevation myocardial infarction. Journal of Cardiovascular Magnetic Resonance. 2022;24:64. doi: 10.1186/s12968-022-00889-7

24. Steding-Ehrenborg K, Arvidsson PM, Töger J, Rydberg M, Heiberg E, Carlsson M, Arheden H. Determinants of kinetic energy of blood flow in the four-chambered heart in athletes and sedentary controls. American Journal of Physiology-Heart and Circulatory Physiology. 2015;310:H113–H122. doi: 10.1152/ajpheart.00544.2015

25. Garg P, Crandon S, Swoboda PP, Fent GJ, Foley JRJ, Chew PG, Brown LAE, Vijayan S, Hassell MECJ, Nijveldt R, et al. Left ventricular blood flow kinetic energy after myocardial infarction - insights from 4D flow cardiovascular magnetic resonance. Journal of cardiovascular magnetic resonance : official journal of the Society for Cardiovascular Magnetic Resonance. 2018;20. doi: 10.1186/s12968-018-0483-6

